# The unequal impact of the COVID-19 pandemic on life expectancy across Chile

**DOI:** 10.1101/2021.12.08.21267475

**Authors:** Gonzalo E. Mena, José Manuel Aburto

## Abstract

**Objectives:** To quantify the effect of the COVID-19 pandemic on life expectancy in Chile categorized by rural and urban, and to correlate life expectancy changes with socioeconomic factors at the municipal level.

**Design:** Retrospective cross-sectional demographic analysis using aggregated data.

**Setting:** Vital and demographic statistics from the national institute of statistics and department of vital statistics of ministry of health.

**Participants:** Aggregated national all-cause death data stratified by year (2000-2020), sex, and municipality.

**Main Outcome measures:** Stratified mortality rates using a Bayesian methodology. With this, we assessed the unequal impact of the pandemic in 2020 on life expectancy across Chilean municipalities for men and women and analyzed previous mortality trends since 2010.

**Results:** Life expectancy declined for both men and women in 2020. Urban areas were the most affected, with males losing 1.89 and females 1.33 in 2020. The strength of the decline in life expectancy correlated with indicators of social deprivation and poverty. Also, inequality in life expectancy between municipalities increased, largely due to excess mortality among the working-age population in socially disadvantaged municipalities.

**Conclusions:** Not only do people in poorer areas live shorter lives, they also have been substantially more affected by the COVID-19 pandemic, leading to increased population health inequalities. Quantifying the impact of the COVID-19 pandemic on life expectancy provides a more comprehensive picture of the toll.

**Strengths and limitations:**

- First study to analyze changes in life expectancy in Chile with small-area resolution.
- We applied a hierarchical Bayesian methodology to estimate life expectancies in the past 20 years.
- We show that COVID-19 led to declines in life expectancy in Chile greater than a year in magnitude. These declines correlated with poverty levels, indicating that socially deprived populations were hit the hardest.
- We also show that inequality in life expectancy between municipalities increased due to excess mortality among the working-age population in socially disadvantaged municipalities.
- The main limitation is that our estimates depend on accurate small-area stratified population estimates. We implemented several estimates and showed that our findings are robust to the choice of them.

## Introduction

Most Latin American countries experienced substantial progress in reducing premature mortality while increasing health standards over the last century and into the first fifteen years of the twenty-first century.^1,2^ But this progress has been reversed, as Latin American countries have been severely affected by the COVID-19 pandemic.^3^ The region became the hotspot of the pandemic in June 2020 and by May 2021 more than one million COVID-19 deaths have been reported.^4,5^

After decades of sustained improvements in life expectancy, leading to levels comparable to low mortality countries, Chile experienced losses in this indicator in 2020 due to increased excess mortality during the COVID-19 pandemic (11 months for women and 1.3 years among men).^6^ While national figures are important and informative, they conceal heterogeneity at the subnational level, which can be substantial. Emerging evidence from Latin American countries suggests that the COVID-19 pandemic has disproportionately affected disadvantaged groups with low socioeconomic status as well as indigenous people, with large regional variation.^7–10^ In Chile’s capital, Santiago, areas with low socioeconomic status experienced poorer health interventions, and substantial excess mortality coupled with higher number of deaths and infection fatality rates at younger ages.^7^ Similarly, municipalities with higher proportions of indigenous population showed higher mortality from COVID-19.^8^ It is unclear, however, what the net effect of increased mortality has been on life expectancy at a more granular level of geography and by population subgroups in Chile.

In this context of persistent and pervasive health inequalities, varied mortality impacts by age and sex, and regional variation, it is imperative to analyze how has life expectancy been affected differently across Chile. Due to the heightened risk to COVID-19 and mortality of disadvantaged populations, most deprived areas may have experienced greater losses in life expectancy, especially among men. Similarly, since rural and urban areas may be affected differently, and mortality increased among young working-age men, we hypothesize that younger age excess mortality will have a substantial effect on life expectancy losses potentially increasing disparities at the municipality level. This hypothesis is supported by evidence from Chile’s capital suggesting that urban and more crowded areas have experienced worse health outcomes during the pandemic.^7,11^ Alternatively, since death rates increased exponentially with age and losses in life expectancy in low mortality countries have been attributed mostly to mortality above age 60,^6^ another hypothesis is that the pandemic in 2020 was such a strong shock that excess mortality differentials decreased, leading to reducing inequalities between municipalities.

This article contributes towards a more comprehensive understanding of the COVID-19 pandemic’s burden on population health by estimating life expectancy across Chilean municipalities by sex using a powerful Bayesian methodology.^12^ We contextualize our results with past trends of progress and disparities in life expectancy, and categorize our findings by urban vs non-urban areas. Our study is a step towards explaining the varied impacts of the pandemic by analyzing trends in life expectancy over age at a more granular level and by correlating life expectancy losses with indicators of poverty in Chile.

## Study Data and Methods

### Data

We used data on births and deaths by age, sex and municipality from publicly available vital statistics.^13^ These data were complemented with official population counts by age (single years of age grom 0 to 89 and collapsed in 90+), sex and municipality from the 2002 and 2017 censuses available from the National Institute of Statistics (INE).^14^ We also used official population projections between 2002 and 2020 centered at the 2017 census.^15^ Unlike censuses themselves, these projections collapsed all ages greater than 80 in one single group. We only observed minor changes in our estimates based on whether the open ended interval started at 80 or 90, but we did observe that life expectancy estimates based on 2017 projections were substantially higher than the ones based on the 2017 census. We explain this by a possible inadequacy of the official projection for later years. Because of this reason, we considered two alternative population estimates for 2017 onwards. The first one assumes that population counts remain fixed for years 2018,2019 and 2020. In the second one, we projected forward the population using the cohort component method^16^ with 2017 as baseline assuming zero migration. We also used census data to classify municipalities as urban or non-urban. (See Supplementary Tables 1-3).^17^ Data on poverty and crowdedness were taken from the CASEN survey by the Chilean Ministry of Social Development and Family.^18^

### Mortality estimation

Age specific death rates for each municipality by sex were estimated implementing a recently developed methodology^12^ based on a hierarchical Bayesian model^19^ using population and death counts.^17^ There are two main advantages to this Bayesian methodology: first, by sharing information across global variables, it is possible to smooth out the noisy estimates that would otherwise be obtained if we relied only on empirical counts. This is important because of the increased likelihood of low death counts on each strata in small municipalities. Second, by appealing to the Bayesian methodology we obtain credible intervals for each of our estimates.

### Life tables

Life tables were calculated using the age specific death rates estimated in the Bayesian procedure following standard techniques.^16^ From these, period life expectancy at birth, temporary life expectancy between ages 20 and 65, and remaining life expectancy at age 65 were subtracted. Life expectancy at birth refers to the average years a cohort of newborns is expected to live given the current mortality conditions. Similarly, life expectancy at age 65 refers to the average years individuals aged 65 are expected to live if they were to experience the current mortality conditions throughout their lives. Given the emerging evidence about how younger age groups below age 65 have also been affected by the pandemic in the context of Chile, we constructed a measure to capture average longevity over working ages through temporary life expectancy. Temporary life expectancy between ages 20 and 65 refers to the average years lived between these ages given prevalent mortality conditions.^20^ For example, if no one were to die between these ages, then the temporary life expectancy would be the full 45 years. To complement our analysis we also consider the probability of not reaching age 65 as an indicator of premature mortality. As a measure of inequality between municipalities we calculated the Gini coefficient of life expectancy across municipalities.^21^ The Gini coefficient is a standard indicator of inequality employed in social sciences. In the context of this paper, the Gini coefficient expresses the degree of inequality in life expectancy across municipalities.

### Limitations

This study had several limitations. First, while Chile’s vital registration is one of the most reliable in Latin America, there are likely to be inaccuracies in mortality registration due to age misreporting and coverage across municipalities, as well as systematic age overstatement.^22^ Delays in recording deaths may lead to incompleteness issues especially in urban areas. Our results on life expectancy declines and mortality inequalities may be considered a lower bound because of these issues. The effect of systematic age overstatement is likely to affect our results too. However, there is no information on what the age pattern of overstatement is during the pandemic. To mitigate these inaccuracies and their effects on our life expectancy estimates, we used a hierarchical Bayesian model that helped to retrieve a reasonable mortality profile across regions. Another limitation is that because of the low number of deaths observed in some municipalities, the degree of uncertainty around the estimates was very high, not allowing us to include them in our analysis with confidence. We excluded municipalities by sex with less than 16,000 people (as per the 2017 census), as we observed that life expectancy estimates were unstable even with our adopted Bayesian methodology. However, we grouped them together and reproduced all results to avoid systematic exclusion. Results were consistent and are shown in Supplementary Figure 2.^17^ Almost all of these were all non-urban municipalities. Some other municipalities were excluded based on a visual inspection of mortality trends that were clearly indicative of coding errors in the mortality database (see Supplementary Figure 1) Despite these limitations, we used the most reliable data for Chile and state-of-the-art methodologies to gauge mortality dynamics across Chile. Finally, our results are limited in that stratified population counts are typically model-based estimates (except at census years), and might be biased. We studied the effect of alternative population estimates in final outcome measures, as described in the Supplement (Figures 3-17).

## Study Results

### Trends in life expectancy at birth and survivorship below age 65

Men and women from both urban and non-urban areas experienced steady increases in life expectancy at birth from 2010 to 2019. Women showed higher life expectancy at birth than men in all groups. In contrast, higher mortality during 2020 led to sharp decreases in life expectancy at birth (Figure 1). Life expectancy among men in urban and non-urban areas declined by 1.89 (1.68,2.09) and 1.66 (1.5,1.8) years, respectively. Among women, life expectancy losses were 1.33 (1.11,1.55) and 1.10 (0.918,1.28), respectively. The magnitude of the decline from 2019 to 2020 offset most gains in life expectancy experienced in the last decade, especially in urban areas. In fact, 68% of the municipalities analyzed ended up with lower life expectancy than in 2015, and this number rose to 75% in urban municipalities.

**Figure 1.**
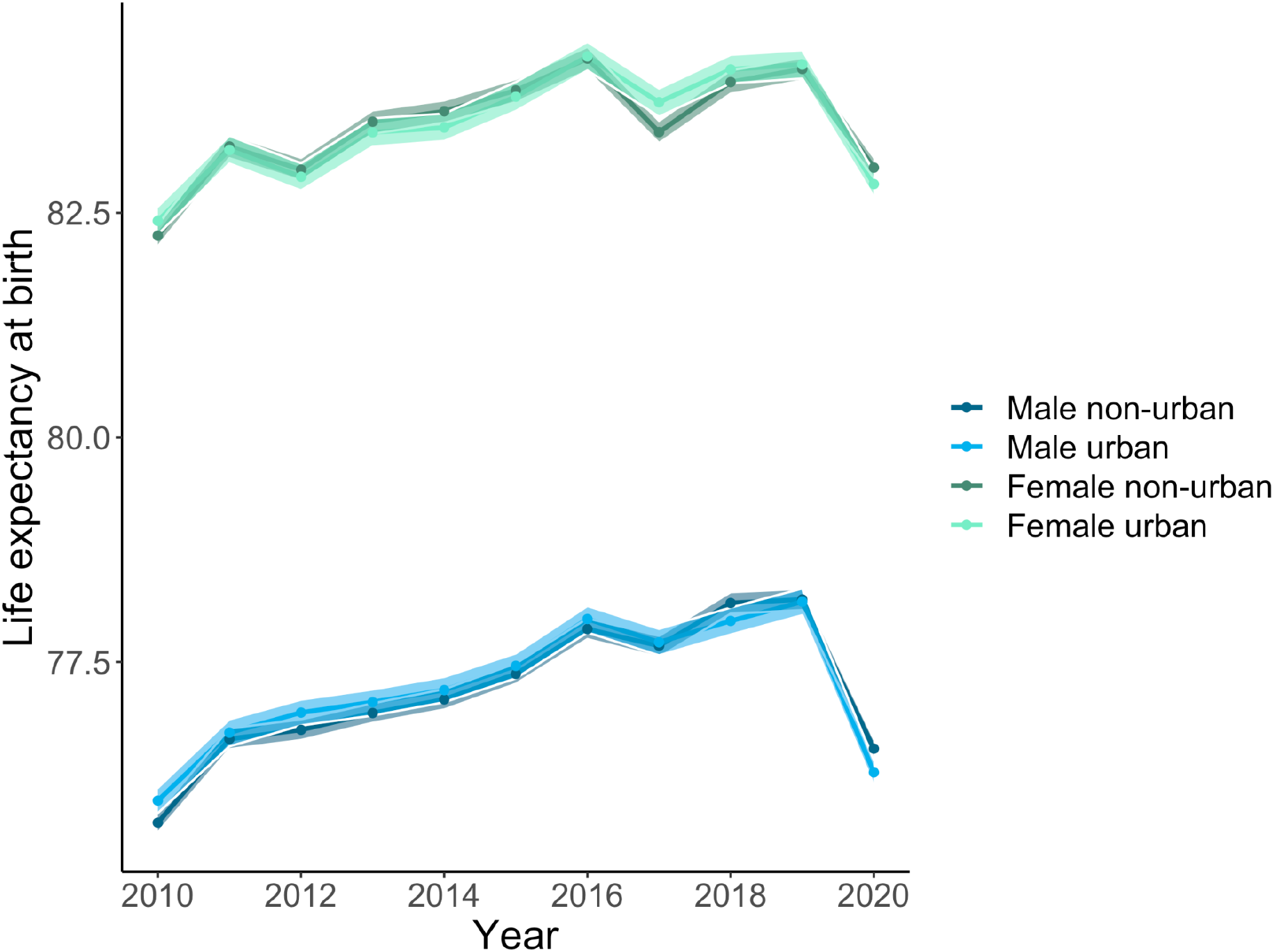
Life expectancy at birth by sex and condition of Urban and Non-urban in Chile. Notes: Solid lines correspond to estimates based on the entire population on each group, with bands indicating 95% credible regions.

Decreases in the probability of surviving to age 65 (Figure 2) indicate that these declines cannot be fully attributed to increased mortality in older age groups only. While mortality above age 65 has been documented as one of the main contributors to declines in life expectancy internationally, substantial increases in mortality below age 65 are apparent in our results, especially among men in urban areas.

**Figure 2.**
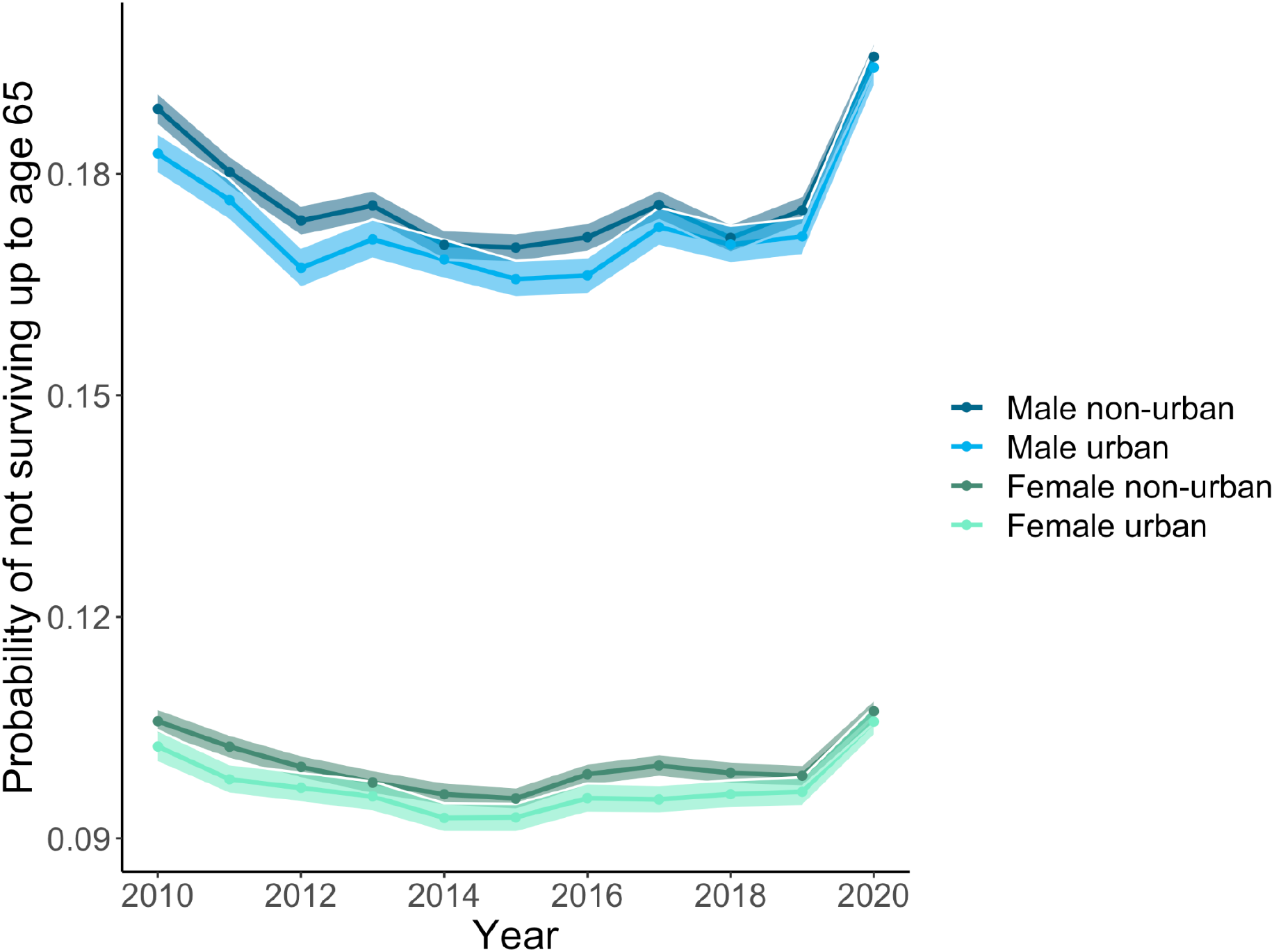
Probabiltiy of not surviving to 65 years by sex and condition of Urban and Non-urban in Chile. Notes: Solid lines correspond to estimates based on the entire population on each group, with bands indicating 95% credible regions.

### Changes in disparities in life expectancy during the COVID-19 pandemic in 2020

Figure 3 shows the year-to-year relative changes of the Gini coefficient as a measure of inequality in life expectancy across municipalities. Panel A refers to life expectancy at birth, panel B to life expectancy between age 20 and 65, and panel C to life expectancy at age 65. From our results it emerges that inequality increased substantially in urban areas from 2019 to 2020 in comparison with previous years, with changes oscillating around 25%. The magnitude of increase is much larger in men and women’s life expectancy between ages 20 and 65 from urban areas (50.9% and 50.6% for men and women respectively). Altogether, these results indicate not only that mortality during 2020 became more unequal, but that this inequality was driven mostly by the younger age group. Supplementary Figures 14-17 show that larger variation in 2020 compared with previous years was driven by lower values of life expectancy.

**Figure 3.**
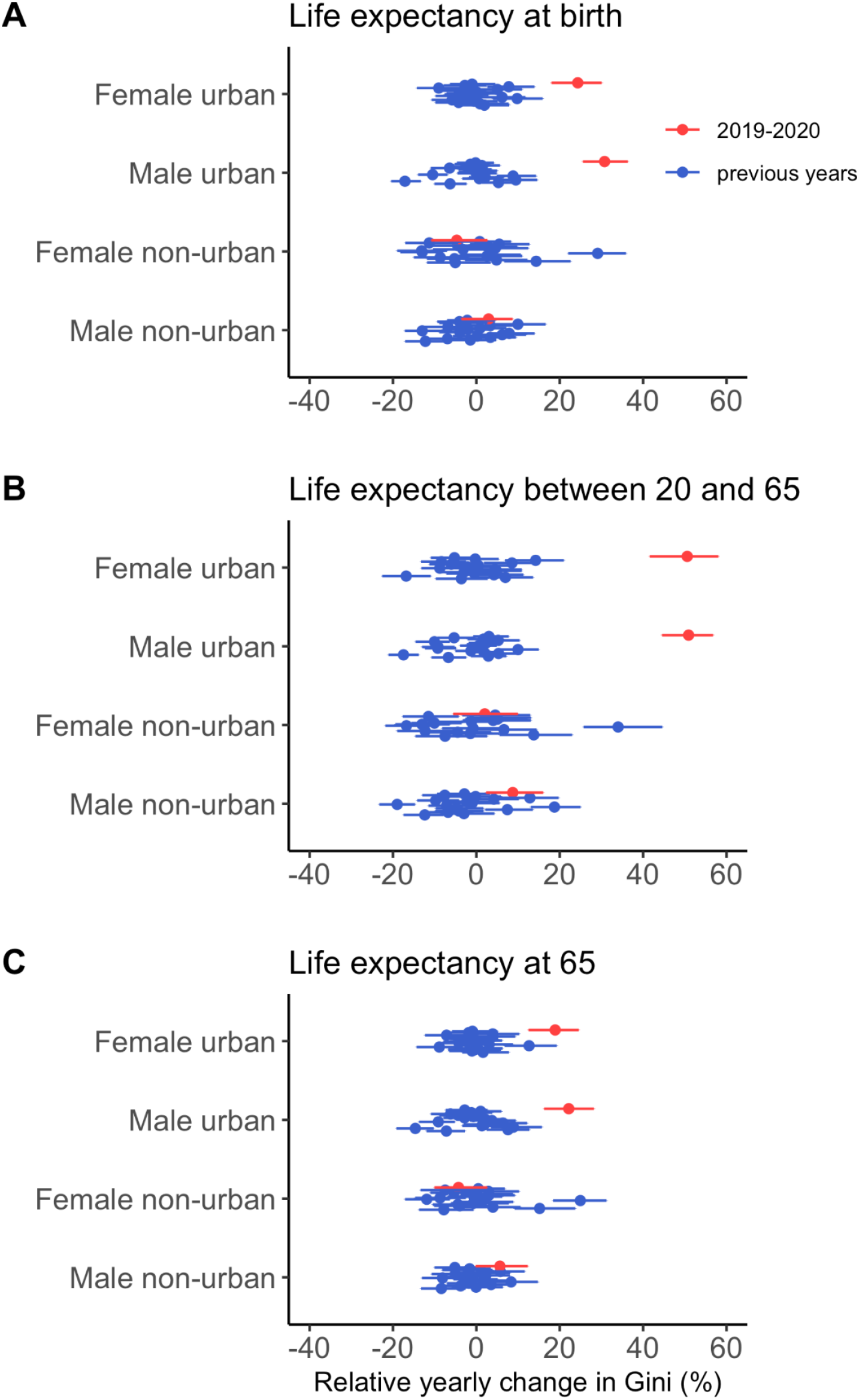
Relative yearly changes in Gini with respect to previous years (starting 2002) for life expectancy at birth (A), average years lived between 20 and 65 (B) and life expectancy at 65.

To better understand the factors driving this spike in inequality, we investigated how declines in life expectancy during 2020 correlated with social deprivation indicators including poverty and crowdedness focusing only on mortality above age 20 across urban areas. Figure 4 shows the relationship between poverty and life expectancy between age 20 ang 65 and life expectancy at age 65. To underscore how the relationship changed in the course of 2020, we stratified the results juxtaposing the previous five years (2015-19) with 2019-20. Results show a strong historical negative correlation between life expectancies in both age groups, sexes and poverty levels. Males in the top poverty decile have a 4.39 expectancy lower life expectancy than in the bottom decile. They also live on average 0.92 less years between 20 and 65, and 2.22 from 65 onwards. For females, these numbers are 2.51, 0.31 and 1.55 years. During 2020, the slope became more negative, suggesting that those municipalities with higher levels of poverty experienced greater losses in life expectancy. This dependency was stronger in the younger age group.

**Figure 4.**
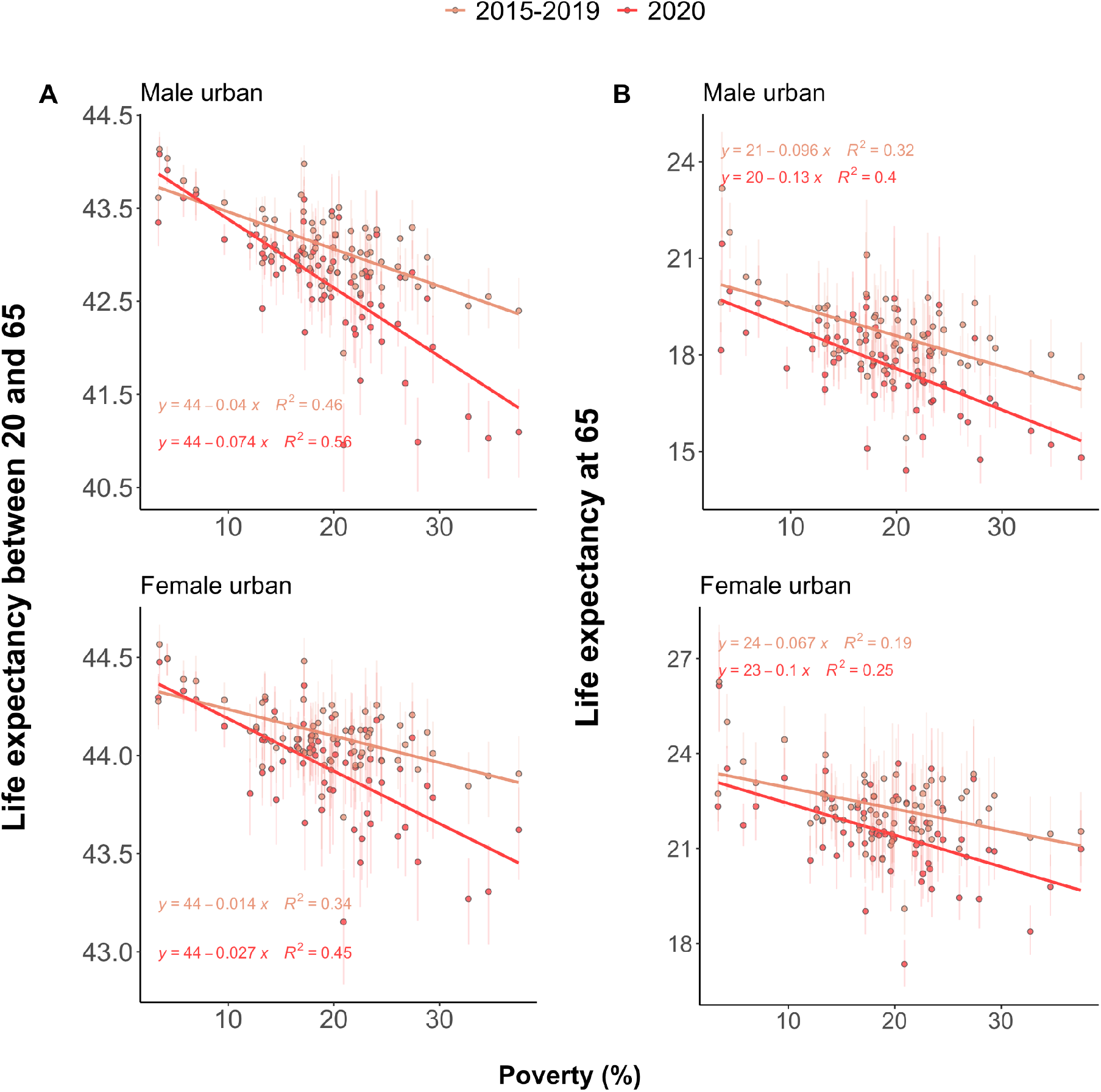
Changes in inequality of mortality in 2020 with respect to recent history were stronger in younger age groups. A Comparison between 2015-2019 and 2020 of the average years lived between 20 and 65, for males and females, as a function of poverty. B same as in A, but with life expectancy at 65.

In contrast, while life expectancy at 65 declined during 2020, this decline was less unequal over the poverty gradient, consistent with the hypothesis that this group contributed less to inequality in changes in life expectancy. To formalize these observations, we performed regression analyses to model the interactions between year and poverty level through varying intercepts and slopes. We only found significant changes in slope for average years lived between 20 and 65. For males, this translated into an additional difference of 0.78 years between the highest and lowest poverty deciles (p=0). For females, this difference was 0.3 (p<0.001)

## Discussion

Urban areas that are exposed to higher poverty or social disadvantages experienced larger losses in life expectancy during the COVID-19 crisis in 2020 in Chile. Our results reveal that losses were unevenly shared across municipales, over age, and by sex, leading to increasing inequality in life expectancy across regions in Chile. Moreover, consistent with previous research on increased mortality at younger ages in 2020 in deprived municipalities in Chile’s capital,^7^ our research shows that working age mortality was one of the main drivers of increasing inequality in life expectancy across Chile.

Analysis of life expectancy in 2020 compared with the previous five years (2015-19) show that poorer urban municipalities suffered a double burden. Not only did they show lower levels of life expectancy but they also experienced greater losses in life expectancy. This is consistent with previous research documenting larger mortality increases for the lower educated groups in Chile’s capital.^23^ Furthermore, when we disaggregate by age groups, we observe that the association between life expectancy for working age individuals (between ages 20 and 65) and levels of poverty became stronger compared to previous years. This is a surprising finding given that previous evidence had documented a positive association between income and life expectancy at retirement.^24^ This suggests that even if the burden of mortality during the COVID-19 crisis has been concentrated at older ages,^25^ contributing substantially to life expectancy declines during 2020,^6^ inequalities in life expectancy were largely driven by increased mortality in working ages at higher levels of poverty. A potential explanation is that the working age population’s availability to work from home and be less exposed to heightened risk of COVID-19 and its consequences varies across municipalities. Deprived populations in Chile’s capital experienced higher fatality rates as a consequence of worse baseline individual health status and to an overwhelmed healthcare system.^7^ Similarly, evidence from the US suggests that those individuals with less availability to work from home had higher death rates compared to those that could afford working from home in 2020.^26^

An open question is whether this sudden increase in inequality amounts to a shock that will be followed by a recovery to pre-pandemic levels, or whether these changes will persist in the long term. Beyond the immediate increase in premature mortality, this is relevant because failing to acknowledge inequalities in mortality may compromise the progressiveness and actuarial fairness of social security and public pension systems in the long term,^27,28^ which could be translated into higher mortality in the future. Similarly, the scars left by the pandemic, including a weak health system, may increase mortality from multiple causes of death. For example, postponed cancer treatments and failure to detect other chronic degenerative diseases timely may lead to lower levels of life expectancy in the long term than it was projected. This highlights the need for accurate and timely data on other causes of death. Future analysis should focus on analyzing the consequences of the COVID-19 pandemic, including multiple causes of death and diseases to study the direct effects from COVID-19 mortality as well as the indirect effects through other pathways of diseases and conditions.^29^ Our research, in this sense, provides a first outlook by focusing on all-cause mortality.

As shown by our results, the case of Chile underscores the dire widening of an already large mortality gap between those living in deprived conditions and those living with higher income during the COVID-19 crisis. Evidence shows that the health consequences of external shocks such a pandemic or an economic crisis are not spread equally across social deprivation levels.^30^ The COVID-19 pandemic reminds us of the ever-present risk of such events, whose cumulative effect may partially explain the ever-existing gaps in mortality. Therefore, the way that this crisis has exposed the vulnerabilities of socially deprived populations is a call to challenge the monolithic view of a country’s demographics in the design of social security systems. New strategies incorporating a public health perspective that considers widening inequalities should be implemented to minimize the effects of the COVID-19 pandemic on the health status of the Chilean population both immediately and in the long term.

## Data Availability

All data is publicly available in www.deis.cl and www.ine.cl

## Acknowledgements

We are grateful to Alberto Palloni for comments on earlier versions of the manuscript, and to Monica Alexander and Ameer Dharamshi for sharing their code related to reference 12.

## Data availability statement

*The data underlying this article are available in The datasets were derived from sources in the public domain:* https://deis.minsal.cl/.

## Author contributions

GM, data curation, software, validation GM and JMA formal analysis, investigation, conceptualisation, methodology, project administration, resources, validation, visualisation, writing (original draft), and writing (review & editing) JMA funding acquisition, supervision

## Ethics approval

This research project does not require ethics approval as it uses only macro data that are freely available online.

## References

1. World Health Organization. The World health report : 2000 : Health systems : improving performance. (World Health Organization, 2000).

2. Alvarez, J.-A., Aburto, J. M. & Canudas-Romo, V. Latin American convergence and divergence towards the mortality profiles of developed countries. Population Studies 74, 75–92 (2020).

3. Castanheira, H. C., Costa Monteiro da Silva, J. H., Del Popolo, F., Guiomar, B. & Saad, P. COVID-19 mortality. Evidence and scenarios. Latin American and Caribbean Demographic Centre (CELADE)-Population Division of ECLAC, United Nations (2021).

4. Dyer, O. Covid-19 hot spots appear across Latin America. BMJ 369, m2182 (2020).

5. Latin America and the Caribbean surpass 1 million COVID deaths - PAHO/WHO | Pan American Health Organization. https://www.paho.org/en/news/21-5-2021-latin-america-and-caribbean-surpass-1-million-covid-deaths.

6. Aburto, J. M. et al. Quantifying impacts of the COVID-19 pandemic through life expectancy losses. medRxiv 2021.03.02.21252772 (2021) doi:10.1101/2021.03.02.21252772.

7. Mena, G. E. et al. Socioeconomic status determines COVID-19 incidence and related mortality in Santiago, Chile. Science (2021) doi:10.1126/science.abg5298.

8. Millalen, P., Nahuelpan, H., Hofflinger, A. & Martinez, E. COVID-19 and Indigenous peoples in Chile: vulnerability to contagion and mortality. AlterNative: An International Journal of Indigenous Peoples 16, 399–402 (2020).

9. Lima, E. et al. Exploring excess of deaths in the context of covid pandemic in selected countries of Latin America. (2020) doi:10.31219/osf.io/xhkp4.

10. Cifuentes, M. P., Rodriguez-Villamizar, L. A., Rojas-Botero, M. L., Alvarez-Moreno, C. A. & Fernández-Niño, J. A. Socioeconomic inequalities associated with mortality for COVID-19 in Colombia: a cohort nationwide study. J Epidemiol Community Health 75, 610–615 (2021).

11. OECD. 1. The COVID-19 crisis in urban and rural areas | OECD Regional Outlook 2021 : Addressing COVID-19 and Moving to Net Zero Greenhouse Gas Emissions | OECD iLibrary. https://www.oecd-ilibrary.org/sites/c734c0fe-en/index.html?itemId=/content/component/c734c0fe-en.

12. Alexander, M., Zagheni, E. & Barbieri, M. A Flexible Bayesian Model for Estimating Subnational Mortality. Demography 54, 2025–2041 (2017).

13. Departamento de Estadísticas Vitales. Departamento de Estadisticas e Información de Salud. https://deis.minsal.cl/.

14. Instituto Nacional de Estadísticas de Chile. Instituto Nacional de Estadísticas de Chile. https://redatam-ine.ine.cl/.

15. INE. Proyecciones de Población. Default http://www.ine.cl/estadisticas/sociales/demografia-y-vitales/proyecciones-de-poblacion.

16. Preston, S. H., Heuveline, P. & Guillot, M. Demography, Measuring and Modeling Population Processes. (Wiley Blackwell, 2001).

17. Health Affairs. Health Affairs supplement. (2021).

18. Ministerio de Desarrollo Social. Observatorio Social - Ministerio de Desarrollo Social y Familia. http://observatorio.ministeriodesarrollosocial.gob.cl/encuesta-casen.

19. Gelman, A., Carlin, J. B., Stern, H. S. & Rubin, D. B. Bayesian Data Analysis. (Chapman and Hall/CRC, 1995). doi:10.1201/9780429258411.

20. Arriaga, E. E. Measuring and Explaining the Change in Life Expectancies. Demography 21, 83–96 (1984).

21. Gini, C. Measurement of Inequality of Incomes. The Economic Journal 31, 124–126 (1921).

22. Palloni, A., Beltrán-Sánchez, H. & Pinto, G. Estimation of older-adult mortality from information distorted by systematic age misreporting. Population Studies 0, 1–18 (2021).

23. Bilal, U., Alfaro, T. & Vives, A. COVID-19 and the worsening of health inequities in Santiago, Chile. Int J Epidemiol dyab007 (2021) doi:10.1093/ije/dyab007.

24. Edwards, R. D. The cost of uncertain life span. J Popul Econ 26, 1485–1522 (2013).

25. Levin, A. T. et al. Assessing the age specificity of infection fatality rates for COVID-19: systematic review, meta-analysis, and public policy implications. Eur J Epidemiol 35, 1123–1138 (2020).

26. Miller, S., Wherry, L. R. & Mazumder, B. Estimated Mortality Increases During The COVID-19 Pandemic By Socioeconomic Status, Race, And Ethnicity: Study examines COVID-19 mortality by socioeconomic status, race, and ethnicity. Health Affairs 40, 1252–1260 (2021).

27. Sanchez-Romero, M., Lee, R. D. & Prskawetz, A. Redistributive effects of different pension systems when longevity varies by socioeconomic status. The Journal of the Economics of Ageing 17, 100259 (2020).

28. Auerbach, A. J. et al. How the Growing Gap in Life Expectancy May Affect Retirement Benefits and Reforms. Geneva Pap Risk Insur Issues Pract 42, 475–499 (2017).

29. Ward, Z. J. et al. Estimating the impact of the COVID-19 pandemic on diagnosis and survival of five cancers in Chile from 2020 to 2030: a simulation-based analysis. The Lancet Oncology 0, (2021).

30. Bambra, C., Riordan, R., Ford, J. & Matthews, F. The COVID-19 pandemic and health inequalities. J Epidemiol Community Health 74, 964–968 (2020).

