## Supplementary text for "The unequal impact of the COVID-19 pandemic on life expectancy across Chile"

#### 1 Municipality classification

Chile is composed by a total of 16 regions. Each region is divided into smaller units, called municipalities. There are a total of 366 municipalities. We classified them as urban or non-urban based on the same criterion as in (*1*), that is, if the following two conditions hold: i) population density greater than 70 people per square kilometer, and ii) the proportion of people living in a urban environment is greater than 88%. We excluded all municipalities having fewer than 16,000 people according to census. In Tables 1 and 2 we show the total number of people and municipalities on urban, non-urban and excluded municipalities. The names of all municipalities and their urbanity status is shown in Table 3. We note that although 147 out of 339 municipalities where excluded, this only signifies a 7% of the population.

To study whether excluding small municipalities would bias our results, we created a super-municipality made by all the excluded. Notably, only two (out of 147) municipalities in this group would have been otherwise categorized as urban (El Quisco, Algarrobo), so it is safe to assume that this super-municipality is a non-urban one. In Fig. 2 we compare time evolution of life expectancy at birth and probability of dying before reaching age 65 (Exhibit 1 and 2 of the main text) for the non-urban municipalities, along with the values for the excluded (mostly non-urban) super-municipality. These are in close agreement.

#### 2 Estimation of mortality rates

We implemented method of (*2*), which consists on a hierarchical Bayesian model for the estimation of age-specific mortality rates on small area setups. The main idea is that by modeling a joint structure for these rates as a function of time and space, it would be possible to smooth out the effect of poor empirical estimates for years/locations where only a few population counts were available. In practice, we found that estimates were reasonable as long as the population of municipalities was reasonably large. We applied the algorithm to all municipalities for each region, and each year between 2002 and 2020, separating by gender (male, female, all). This gave a total of $16 \times 3$ algorithm runs. For each a run, we obtained a total of 3,000 Monte Carlo samples that we used for computing credible intervals. Additionally, we ran the algorithm to compute mortality rates for each region, and for the totality of urban and non-urban municipalities, as necessary. In all cases, we estimated mortality rates based on 5 years intervals, up to age 80+ (see below for a discussion of the cutoff age).

We excluded from our analyses some municipalities/years based on the visual inspection of total deaths per year. A cluster of 6 municipalities appeared to have corrupted data in the years surrounding 2004. Those are shown in Fig. 1.

#### 3 Regressions

#### 4 Sensitivity analyses

Since deaths are revealed to us in full detail, and because Chilean death recording system is reliable (*3*), the main source of corruption in mortality rates should stem from possible biases in population estimates. We explored what was the impact of different ways using population estimates in constructing the life tables, and used a number of several alternative estimates to re-create the results shown in the main text. These are explained below.

**Improving official projections**

For year specific population counts between 2002 and 2020, we used the official population projections provided by the national institute of statistics, available at the municipality level and with resolution of years. These are made with simple interpolation and extrapolation methods as described in (*4*). However, we found that these projections were often inconsistent, mostly from 2017 on. Therefore, we considered two alternative estimates in addition to official projections, that only differed from official estimates starting 2017. For one estimate we used the official census counts at 2017 for years 2018, 2019 and 2020. The second estimate corresponds to the cohort component projection method, where we used births in 2017 (the only available) and deaths in 2018, 2019, 2020 to infer municipality and age specific population counts after 2017. In Fig. 5 we show comparisons between resulting estimates. We observe that indeed they produce different estimates, and differences between methods increase for later years. Notably, estimates based on official projections deviate wildly from other in some municipalities, indicating a possible lack of accuracy. In particular, we should expect that estimations based on projections at census year 2017 should be similar to the ones provided by our alternative estimates.

**Maximum age**

Another source of bias is given by cutoff age used when turning age-specific mortality rates into life expectancy estimates. Official census information (2002,2017) contains age-specific population counts for each municipality and gender, up to age 90. However, official census projections collapses all ages above 80 into one group. In Fig. 5A we compare results with the 80 and 90 cutoff, using official census data (only years 2002 and 2017), We observe that the 90 cutoff leads to consistently slightly higher life expectancies, with a difference that appears higher for older ages. Importantly, in 5B,C we also include other estimates, for reference. We observe large discrepancies in year 2017 when comparing official census and official projections. Once more, this is an indication that official projections are not accurate, as they become inconsistent in 2017 (i.e., official projections in year 2017 are far from official census in the same year).

**Main results with alternative estimates** In the main text we have used the cohort survival projection method. Here, we present results using the other two alternative methods. Figs. 5 and 6 correspond to Exhibits 1 and 2 in the main text, respectively. Figs. 7 and 8 complement Exhibit 3, and likewise, Figs. 9 and 10 complement Exhibit 4.

### 5 Additional results

We provide additional figures that supplement exhibits in the main text. In Fig. 11 we show histograms of the life expectancy (with each sample representing a municipality) at even years. We observe that a left tail appears during 2020 (mostly in urban setups) indicative of the unequal impact of COVID-19 in some municipalities. Fig. 12 supplements Exhibit 3 in the main text, but showing the entire Gini time series, and not only the year-to-year differences. A clear abrupt increase is observed during 2020. Interestingly, a consistent temporal drop in Gini is observed between 2002-2019 (with the exception of 2010, when an earthquake caused hundreds of casualties localized in space), for life expectancies between 20-40. Finally, Fig.13 supplements Exhibit 4 by showing the relation between life expectancy and poverty in non-urban municipalities. No clear consistent pattern is observed. Also, in Fig. 14 we show the corresponding decreases of life expectancy over time as a function of poverty, in urban and non-urban setups. This figure is complemented by Fig. 15, which shows an even stronger correlation when using crowdedness as covariate, and Figs. 16 and 17, which show sensitivity of Fig. 14 to changes in the projection methodology.

| Region | Urban | Rural | Excluded | Total |
|---|---|---|---|---|
| Tarapaca | 299843 | 0 | 30715 | 330558 |
| Antofagasta | 0 | 552790 | 54744 | 607534 |
| Atacama | 448784 | 251371 | 57431 | 757586 |
| Coquimbo | 880647 | 787549 | 139030 | 1807226 |
| Valparaíso | 0 | 223516 | 62652 | 286168 |
| O'Higgins | 275211 | 477699 | 161645 | 914555 |
| Maule | 369493 | 559301 | 116156 | 1044950 |
| Biobio | 946952 | 504405 | 105448 | 1556805 |
| La Araucanía | 282415 | 522213 | 140985 | 945613 |
| Los Lagos | 407362 | 262009 | 159337 | 828708 |
| Aysen | 0 | 81777 | 20233 | 102010 |
| Magallanes | 0 | 153069 | 12304 | 165373 |
| Metropolitana | 6273435 | 809613 | 29760 | 7112808 |
| Los Ríos | 166080 | 181799 | 36958 | 384837 |
| Arica y Parinacota | 0 | 221364 | 4704 | 226068 |
| Nuble | 215646 | 152749 | 100611 | 469006 |
| Chile | 10565868 | 5741224 | 1232713 | 17539805 |

Table 1: Total populations for each region for each strata (urban, rural) in our design.

### References


1. J. Berdegué, E. Jara, F. Modrego, X. Sanclemente, A. Schejtman, *Rimisp, Santiago* (2009).

2. M. Alexander, E. Zagheni, M. Barbieri, *Demography* **54**, 2025 (2017).

3. G. E. Mena, *et al.*, *Science* **372** (2021).

4. I. N. de Estadísticas, Estimaciones y proyecciones de la población de chile 2002-2035 a nivel comunal. documento metodológico (2019 [Online].).

| | | Urban | Rural | Excluded | Total |
|---|---|---|---|---|---|
| Region | Tarapaca | 2 | 0 | 5 | 7 |
| | Antofagasta | 0 | 3 | 6 | 9 |
| | Atacama | 0 | 3 | 6 | 9 |
| | Coquimbo | 2 | 6 | 7 | 15 |
| | Valparaíso | 9 | 15 | 14 | 38 |
| | O'Higgins | 2 | 14 | 17 | 33 |
| | Maule | 2 | 15 | 13 | 30 |
| | Biobio | 9 | 12 | 12 | 33 |
| | La Araucanía | 1 | 16 | 14 | 31 |
| | Los Lagos | 2 | 9 | 19 | 30 |
| | Aysen | 0 | 2 | 6 | 8 |
| | Magallanes | 0 | 2 | 6 | 8 |
| | Metropolitana | 36 | 13 | 3 | 52 |
| | Los Ríos | 1 | 7 | 4 | 12 |
| | Arica y Parinacota | 0 | 1 | 3 | 4 |
| | Nuble | 2 | 6 | 12 | 20 |
| | Chile | 68 | 124 | 147 | 339 |

Table 2: Number of municipalities for each strata (urban, rural) in our design, for each region.

| Region | Municipalities |
|---|---|
| Tarapaca | Iquique, Alto Hospicio, Pozo Almonte, Camina, Colchane, Huara, Pica |
| Antofagasta | Antofagasta, Calama, Tocopilla, Mejillones, Sierra Gorda, Taltal, Ollague, San Pedro de Atacama, Maria Elena |
| Atacama | Copiapo, Caldera, Vallenar, Tierra Amarilla, Chanaral, Diego de Almagro, Alto del Carmen, Freirina, Huasco |
| Coquimbo | La Serena, Coquimbo, Vicuna, Illapel, Los Vilos, Salamanca, Ovalle, Monte Patria, Andacollo, La Higuera, Paiguano, Canela, Combarbala, Punitaqui, Rio Hurtado. |
| Valparaíso | Valparaiso, Concon, Calera, La Cruz, San Antonio, Cartagena, San Felipe, Quilpue, Villa Alemana, Casablanca, Puchuncavi, Quintero, Vina del Mar, Los Andes, San Esteban, La Ligua, Cabildo, Quillota, Hijuelas, Nogales, Llaillay, Putaendo, Limache, Olmue, Juan Fernandez, Isla de Pascua, Calle Larga, Rinconada, Papudo, Petorca, Zapallar, Algarrobo, El Quisco, El Tabo, Santo Domingo, Catemu, Panquehue, Santa Maria |
| O'Higgins | Rancagua, Graneros, Coltauco, Donihue, Las Cabras, Machali, Mostazal, Pichidegua, Rengo, Requinoa, San Vicente, Pichilemu, San Fernando, Chimbarongo, Nancagua, Santa Cruz, Codegua, Coinco, Malloa, Olivar, Peumo, Quinta de Tilcoco, La Estrella, Litueche, Marchihue, Navidad, Paredones, Chepica, Lolol, Palmilla, Peralillo, Placilla, Pumanque |
| Maule | Talca, Curico, Constitucion, Maule, San Clemente, Cauquenes, Molina, Sagrada Familia, Teno, Linares, Colbun, Longavi, Parral, Retiro, San Javier, Villa Alegre, Yerbas Buenas Curepto, Empedrado, Pelarco, Pencahue, Rio Claro, San Rafael, Chanco, Pelluhue, Hualane, Licanten, Rauco, Romeral, Vichuquen |
| Biobio | Concepcion, Coronel, Chiguayante, Lota, Penco, San Pedro de la Paz, Talcahuano, Tome, Hualpen, Hualqui, Lebu, Arauco, Canete, Curanilahue, Los Alamos, Los Angeles, Cabrero, Laja, Mulchen, Nacimiento, Yumbel Florida, Santa Juana, Contulmo, Tirua, Antuco, Negrete, Quilaco, Quilleco, San Rosendo, Santa Barbara, Tucapel, Alto Biobio |
| La Araucanía | Temuco, Carahue, Cunco, Freire, Lautaro, Loncoche, Nueva Imperial, Padre Las Casas, Pitrufquen, Pucon, Vilcun, Villarrica, Angol, Collipulli, Curacautin, Traiguen, Victoria, Curarrehue, Galvarino, Gorbea, Melipeuco, Perquenco, Saavedra, Teodoro Schmidt, Tolten, Ercilla, Lonquimay, Los Sauces, Lumaco, Puren, Renaico |
| Los Lagos | Puerto Montt, Osorno, Calbuco, Frutillar, Los Muermos, Llanquihue, Puerto Varas, Castro, Ancud, Quellon, Purranque, Cochamo, Fresia, Maullin, Chonchi, Curaco de Velez, Dalcahue, Puqueldon, Queilen, Quemchi, Quinchao, Puerto Octay, Puyehue, Rio Negro, San Juan de la Costa, San Pablo, Chaiten, Futaleufu, Hualaihue, Palena |
| Aysen | Coyhaique, Aysén Lago Verde, Cisnes, Guaitecas, Cochrane, Chile Chico, Rio Ibanez |
| Magallanes | Punta Arenas, Natales Laguna Blanca, San Gregorio, Cabo de Hornos, Porvenir, Primavera, To[illegible] |
| Metropolitana | Santiago,Cerrillos, Cerro Navia, Conchali, El Bosque, Estacion Central, Huechuraba, Independencia, La Cisterna, La Florida, La Granja, La Pintana, La Reina, Las Condes, Lo Barnechea, Lo Espejo, Lo Prado, Macul, Maipu, Nunoa, Pedro Aguirre Cerda, Penalolen, Providencia, Pudahuel, Quilicura, Quinta Normal, Recoleta, Renca, San Joaquin, San Miguel, San Ramon, Vitacura, Puente Alto, San Bernardo, Padre Hurtado,Penaflor, Pirque, San Jose de Maipo, Colina, Lampa, Tiltil, Buin, Calera de Tango, Paine, Melipilla, Curacavi, Talagante, El Monte, Isla de Maipo, Alhue, Maria Pinto, San Pedro |
| Los Ríos | Valdivia, Lanco, Los Lagos, Mariquina, Paillaco, Panguipulli, La Union, Rio Bueno, Corral, Mafil, Futrono, Lago Ranco |
| Arica y Parinacota | Arica Camarones, Putre, General Lagos |
| Nuble | Chillan, Chillan Viejo, Bulnes, Quillon, San Ignacio, Yungay, San Carlos, Coihueco, El Carmen, Pemuco, Pinto, Quirihue, Cobquecura, . Coelemu, Ninhue, Portezuelo, Ranquil, Treguaco, Niquen, San Fabian |



Table 3: Names of all urban (red), rural (blue) and excluded (black) municipalities of each region.

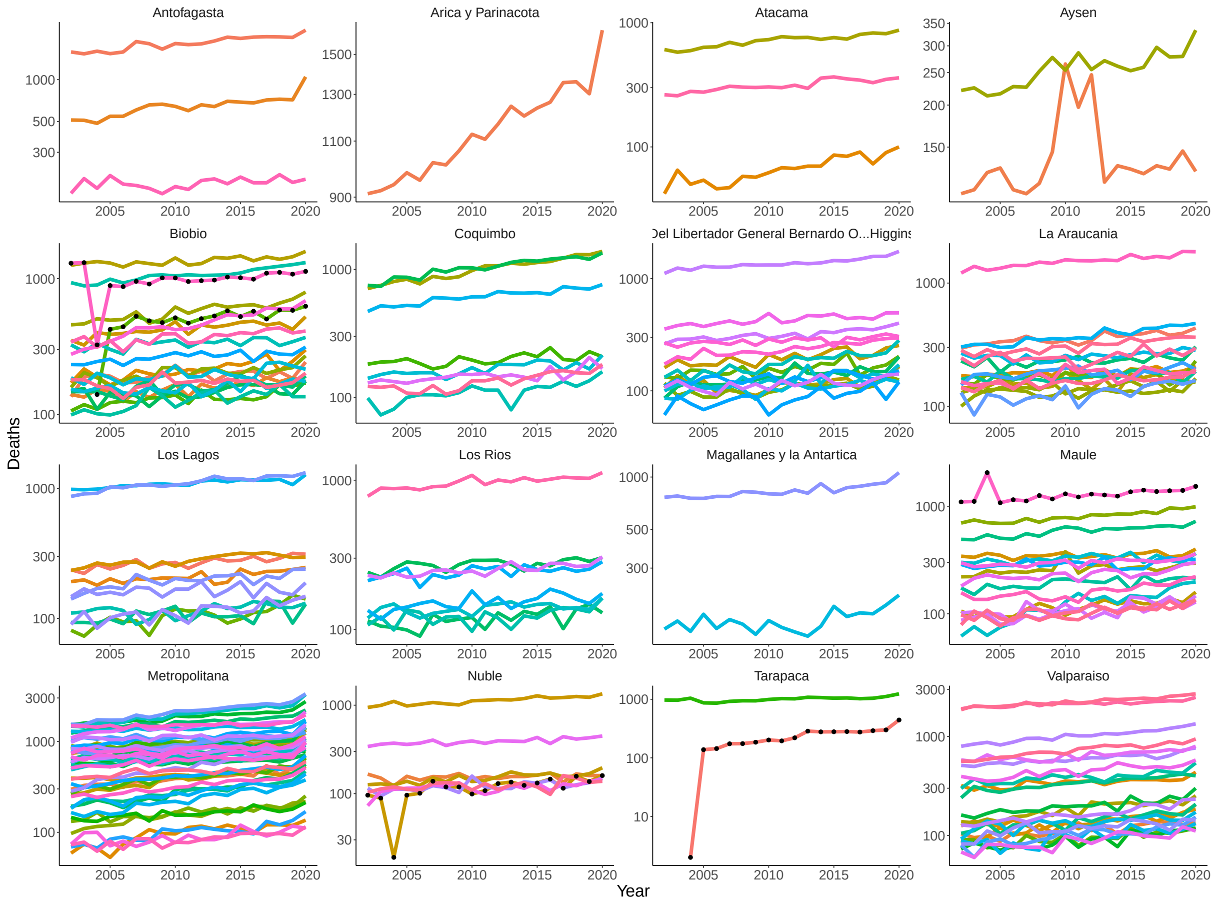


Figure 1: Yearly deaths for each municipality (colored lines) grouped by region (different plots). Lines that are also dotted are the ones for which anomalies existed in recording, leading to sudden drops and/or increases around 2004, presumably due to coding errors. These were excluded in the neighboring years (Talcahuano, Hualpén, Diego de Almagro, Talca, Alto Hospicio, Chillán Viejo).

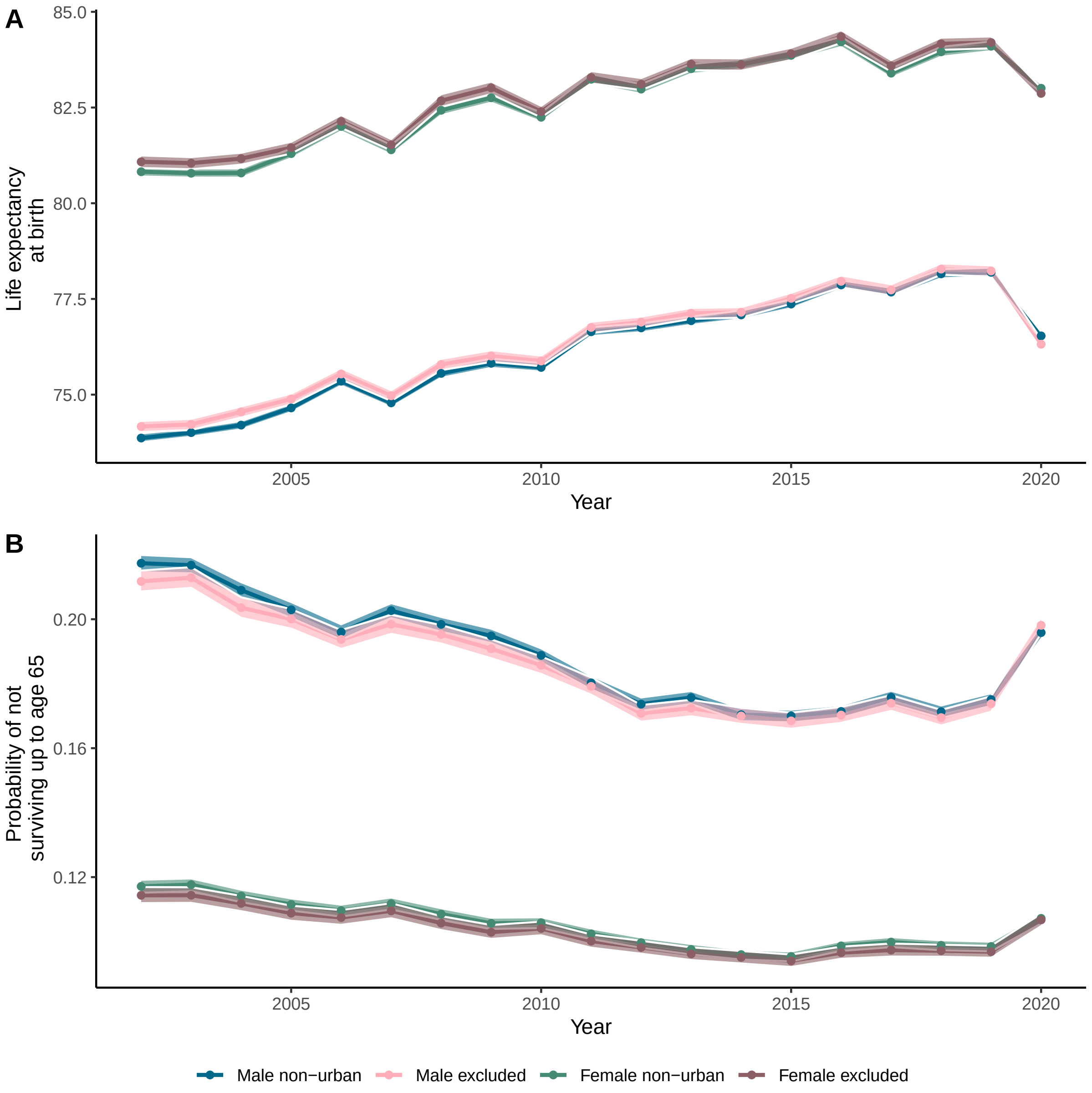


Figure 2: **A**. Time evolution of life expectancy, including the excluded municipalities collapsed as a super-municipality. **B**. Same as **A**, but with likelihood of dying before reaching 65.

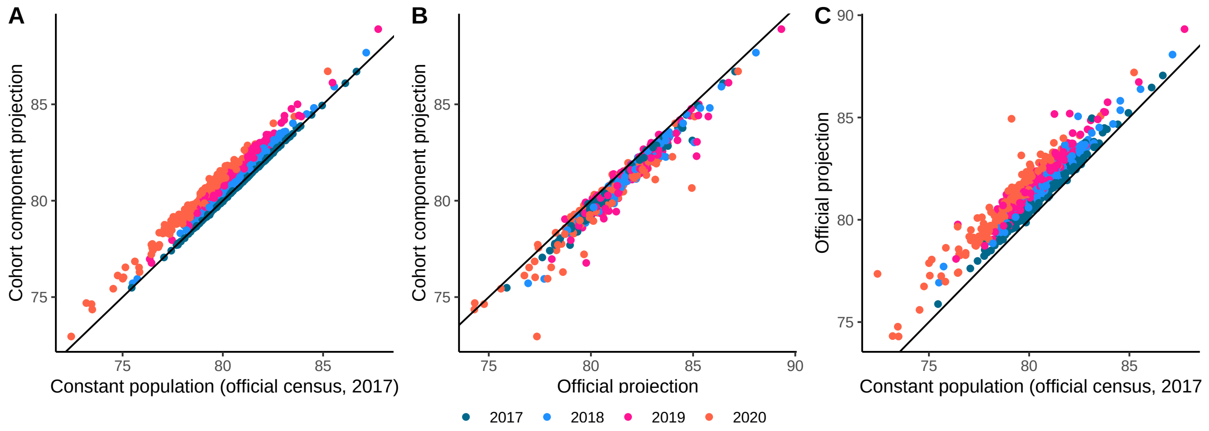


Figure 3: Comparison of various life expectancy estimates, for years 2017-2020. All of these use 80 as cutoff age for population counts. In **A** we compare cohort survival projection with the one that makes the population constant from 2017 on. In **B** we compare official projections with cohort survival projection. In **C** we compare official projection with the one that has constant population.

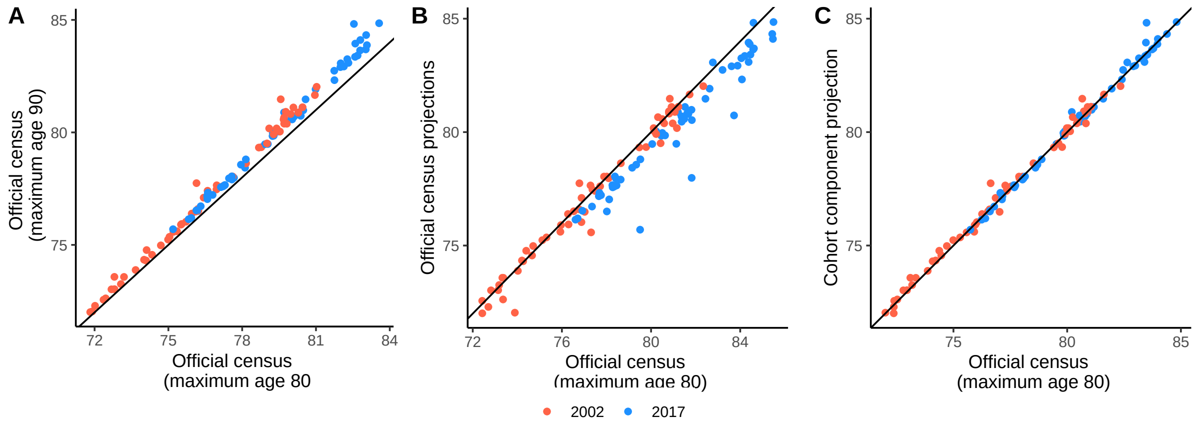


Figure 4: Comparison of several life expectancy estimates, only for census years (2002, 2017). In **A** we compare estimates based on census data but different age cutoffs. When using 90 as cutoff, life expectancies appear slightly higher. In **B** we compare the official census data with 80 cutoff with official projections in that year. We note that discrepancies become more significant in year 2017, indicating the need for an alternative methodology. In **C** we compare official census (80 as cutoff age) with our cohort survival projection method. They are in close agreement, as they are both based on official census data, and not projections.

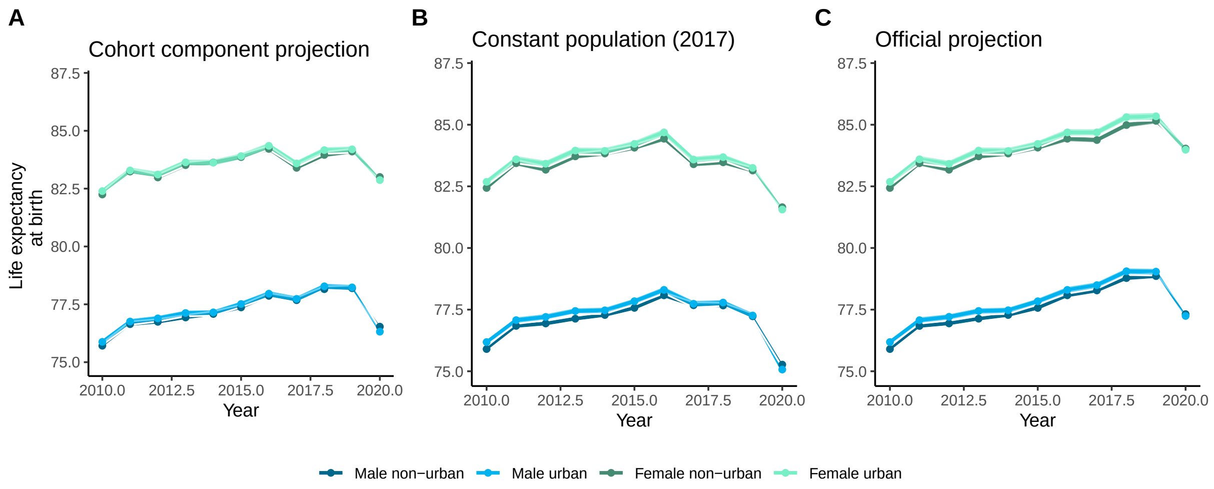


Figure 5: Time evolution of life expectancy, using our three estimators, Exhibit 1 in main text coincides with **A**.

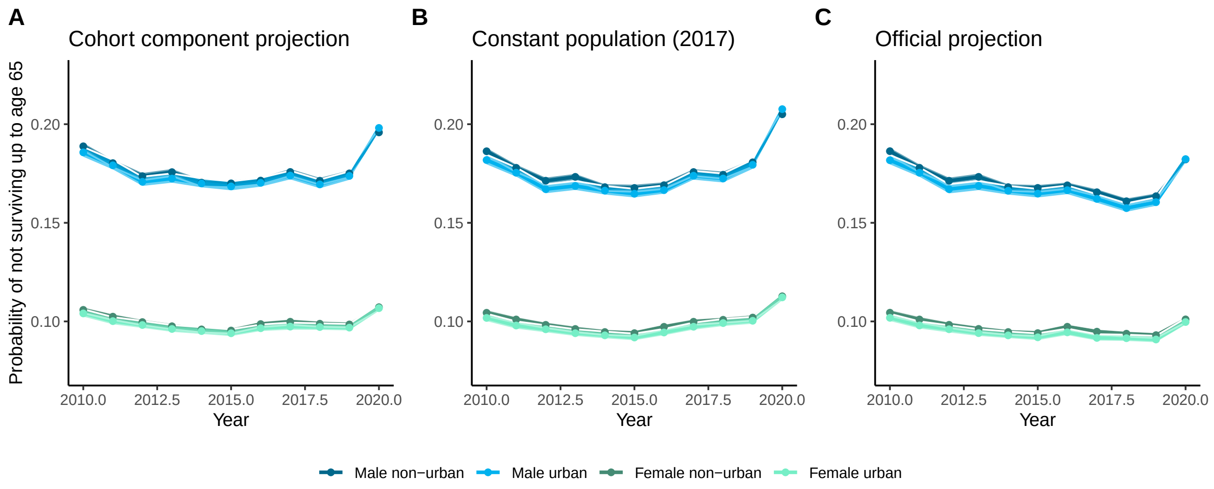


Figure 6: Time evolution probability of not surviving up to 65 years, using our three estimators. Exhibit 2 in main text coincides with **A**.

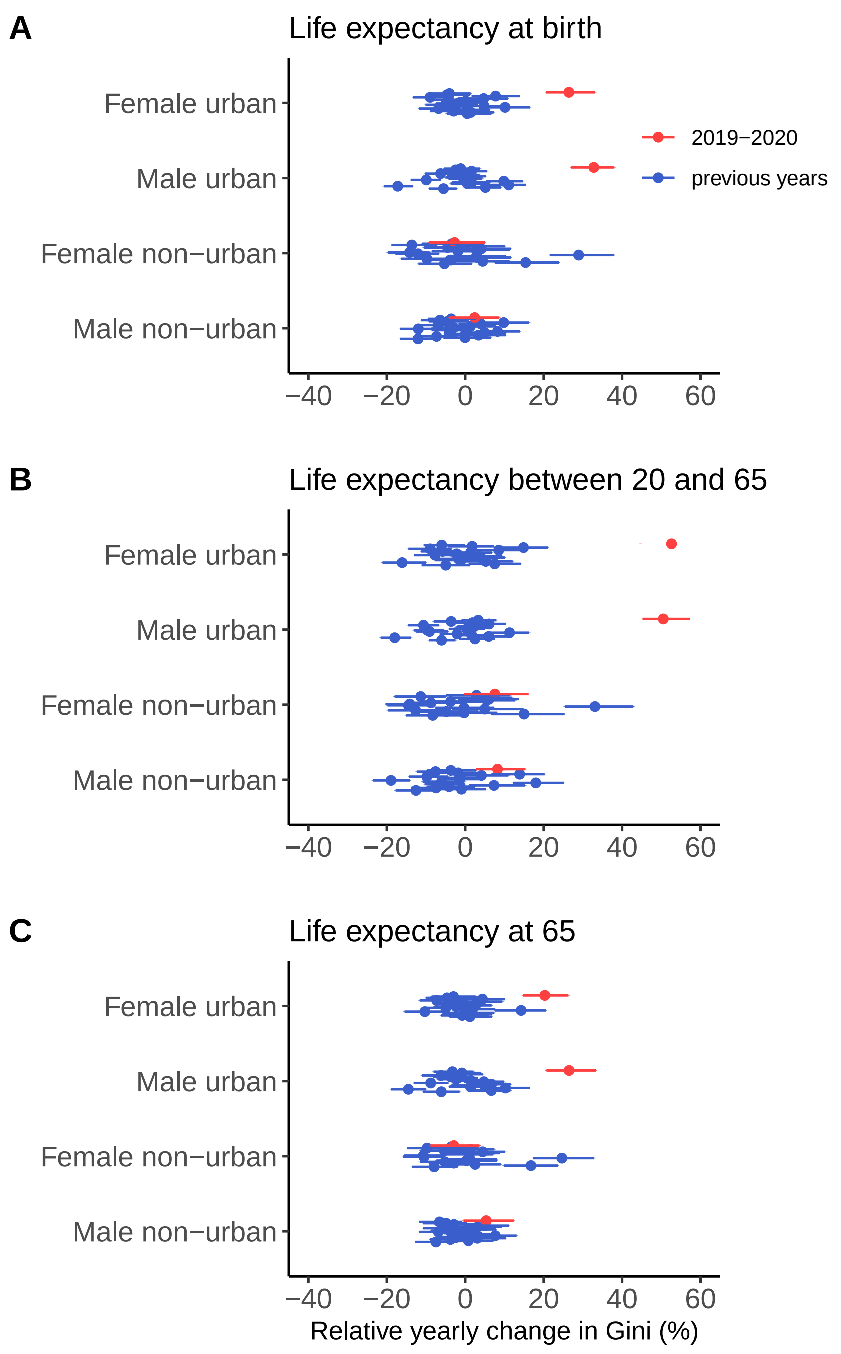


Figure 7: Year-to-year relative changes in Gini, where we have assumed that population after 2017 remained constant (equal to the one provided by census). Bars represent 75% credible intervals. This figure supplements Exhibit 3 in the main text.

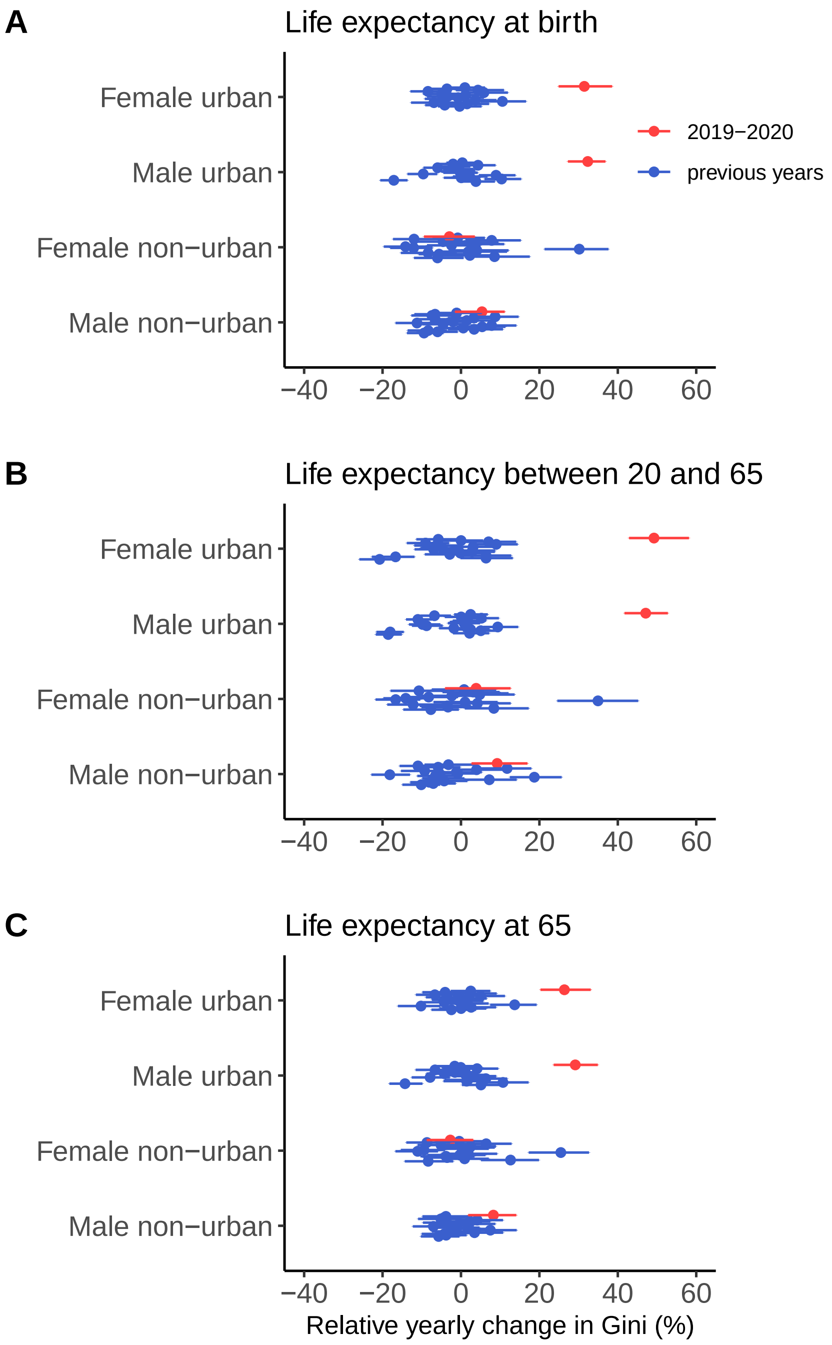


Figure 8: Year-to-year relative changes in Gini, where we have used the official census projections. Bars represent 75% credible intervals. This figure supplements Exhibit 3 in the main text.

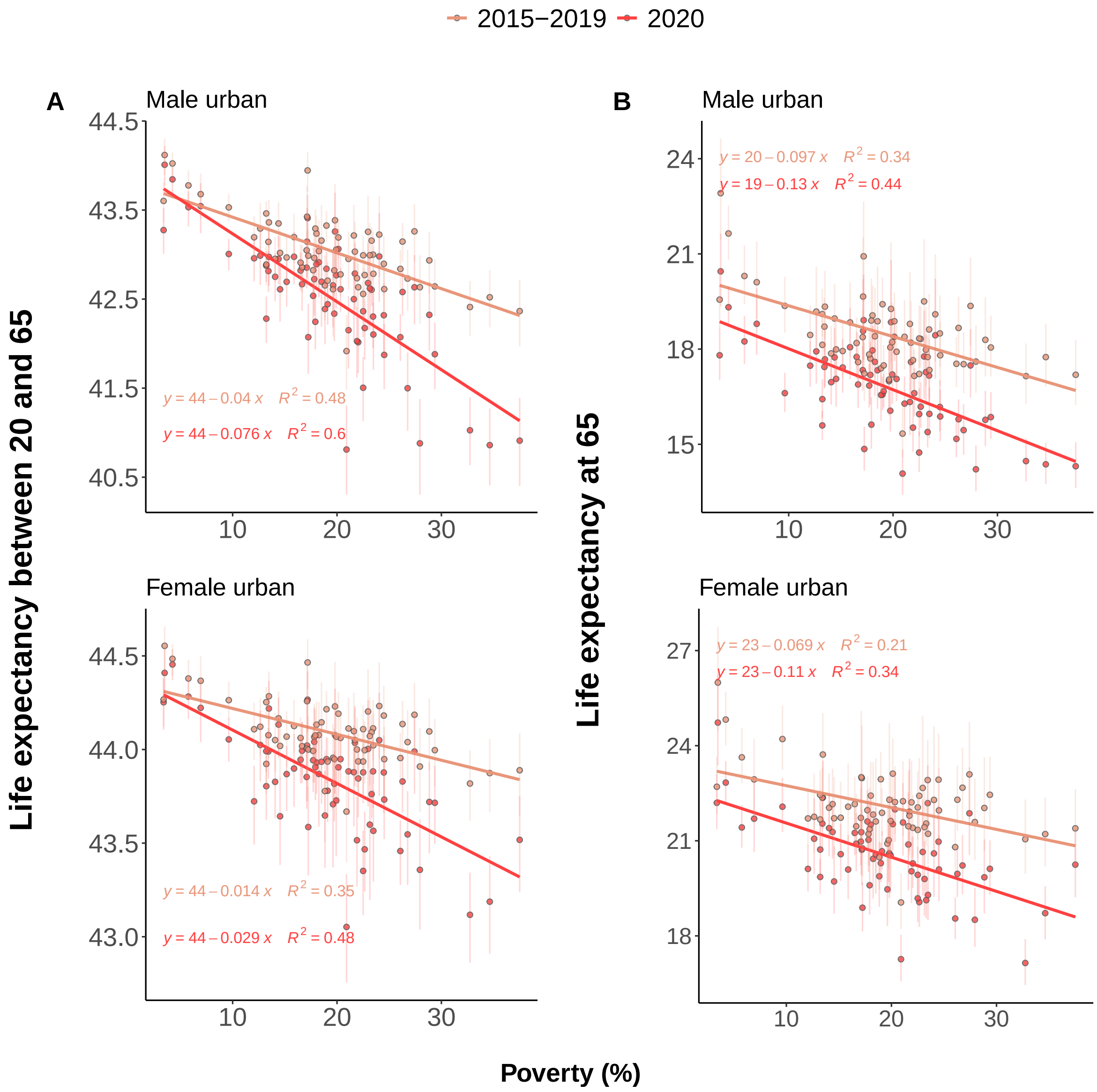


Figure 9: **A** Life expectancy between 20 and 65 and **B** and life expectancy at 65 as a function of poverty and gender, for urban municipalities. Bars represent 95% credible intervals. These estimates are based on the method that fixed population counts at values in 2017 for years 2017, 2018, 2019 and 2020, and may be compared with Exhibit 4 in the main text.

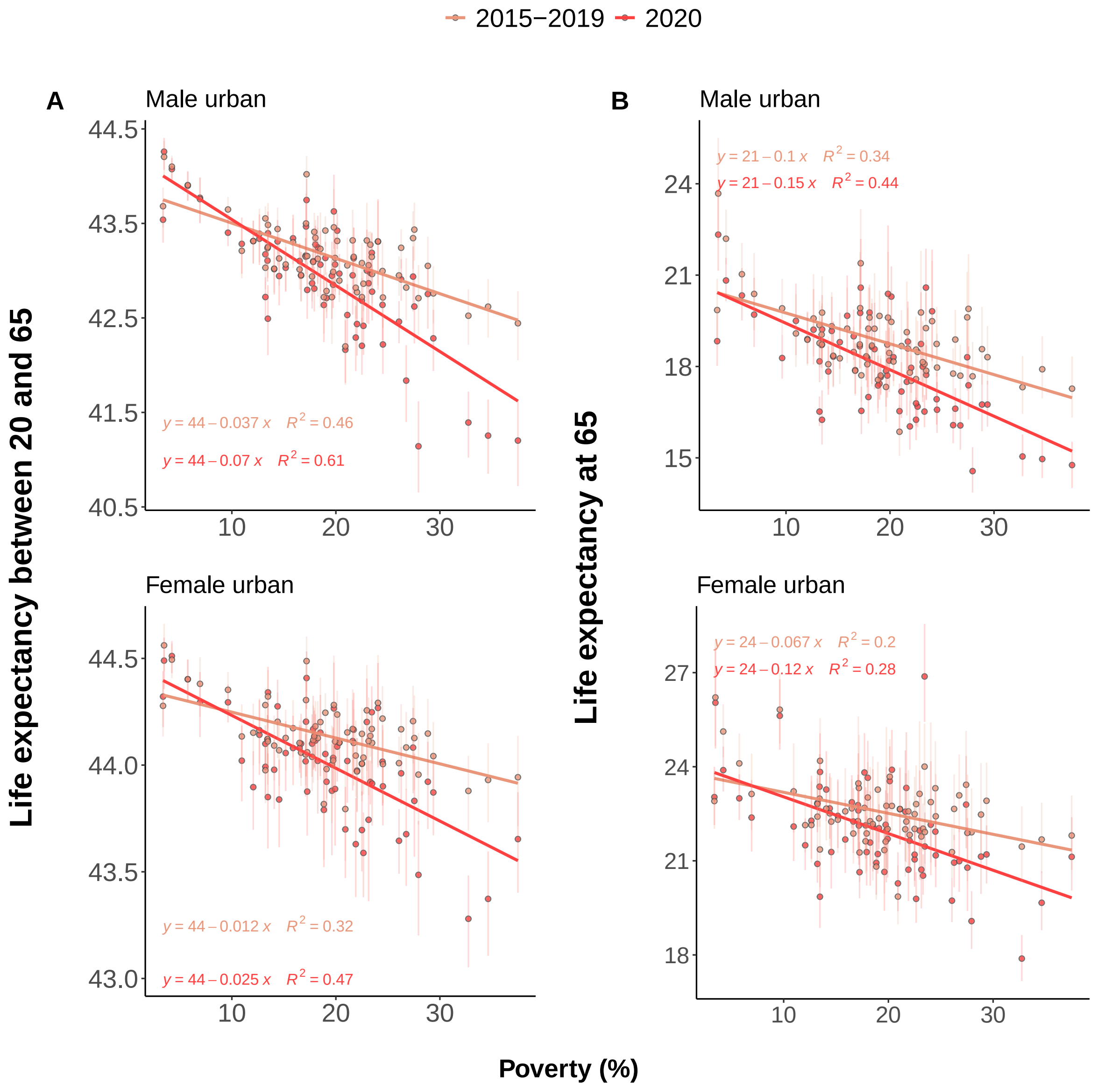


Figure 10: **A** Life expectancy between 20 and 65 and **B** and life expectancy at 65 as a function of poverty and gender, for urban municipalities. These estimates are based on the official census projections and may be compared with Exhibit 4 in the main text.

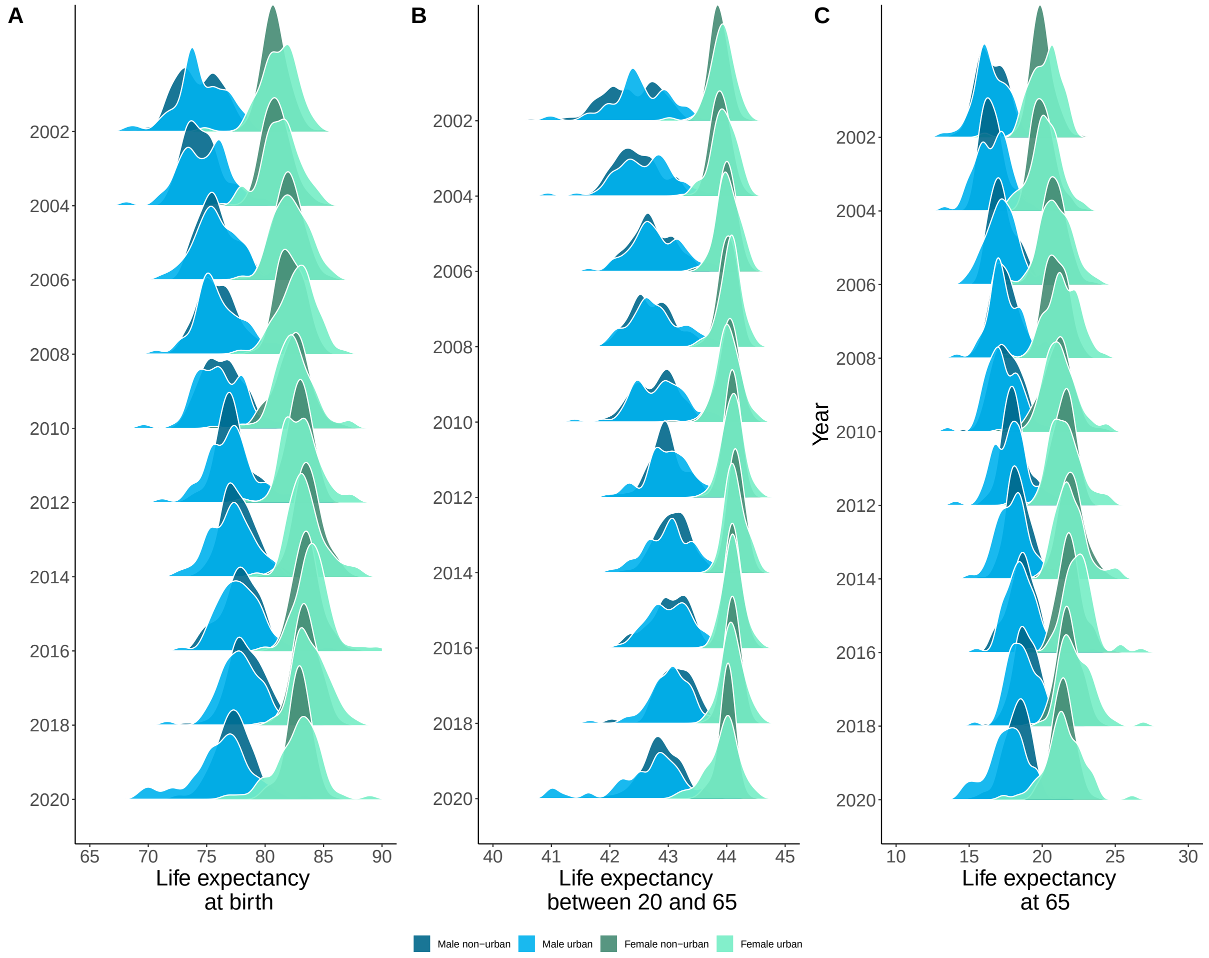


Figure 11: Histograms of life expectancies over time, for male/female and urban/non-urban settings. These histograms are made by taking each combination of gender as municipality as a sample. We note that a left tail appears during 2020 for urban municipalities

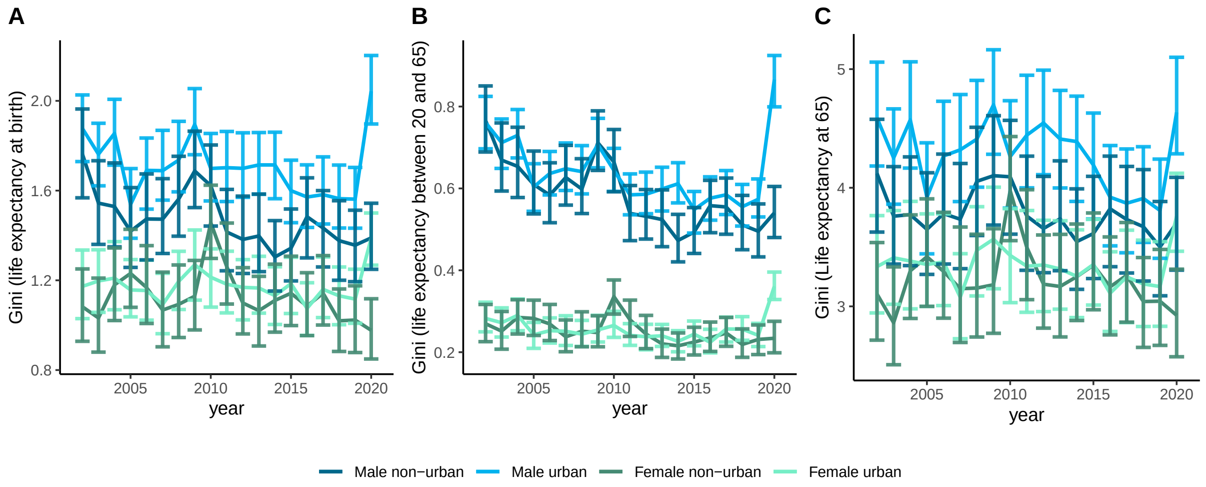


Figure 12: Time evolution of Gini. This plot supplements Exhibit 3, where only year-to-year differences are shown. Bars represent 95% credible intervals.

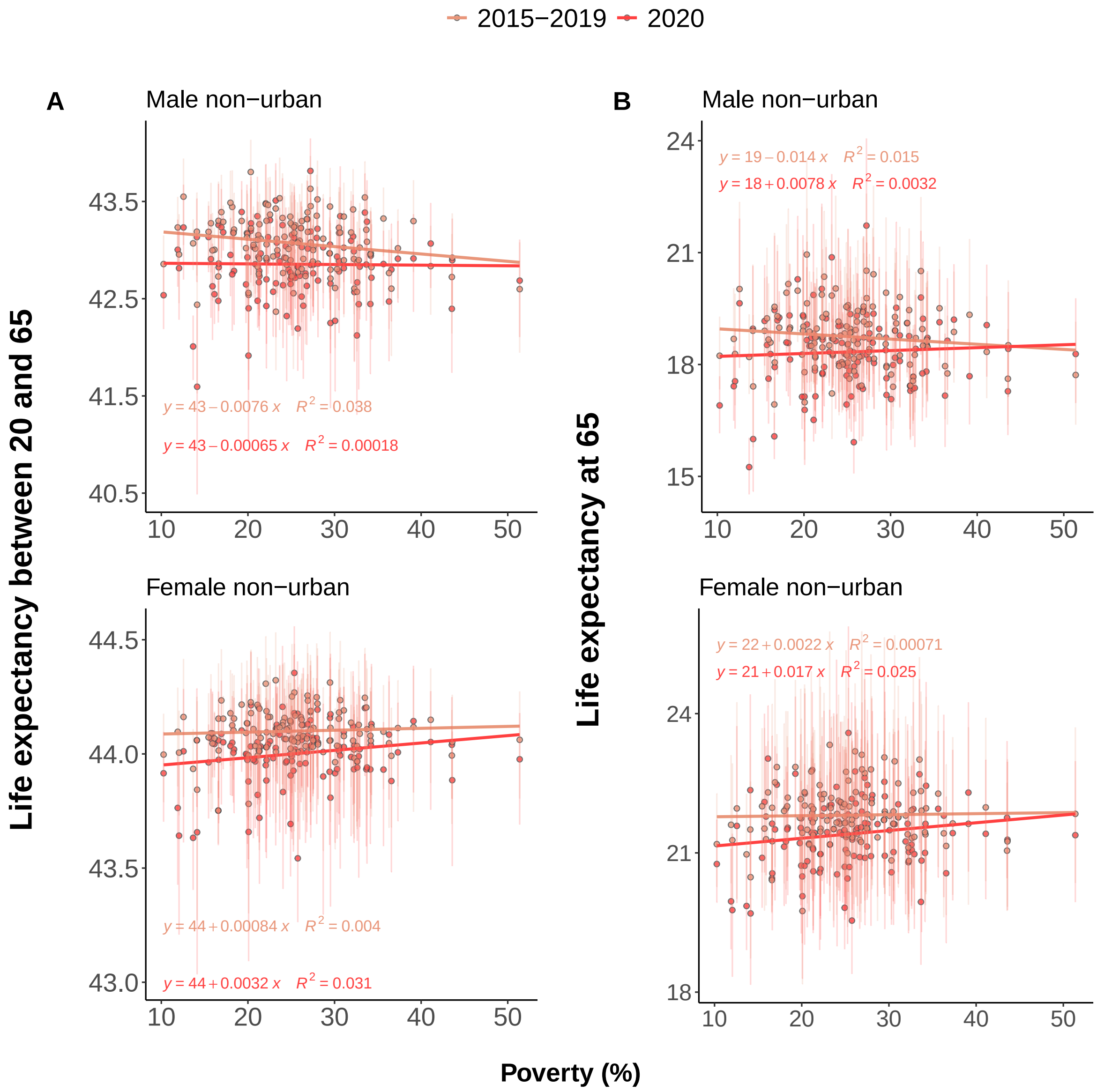


Figure 13: **A** Life expectancy between 20 and 65 and **B** and life expectancy at 65 as a function of poverty and gender, for non-urban municipalities. These are similar to results in Exhibit 4 in the main text, but correlations vanish when focusing on non-urban municipalities.

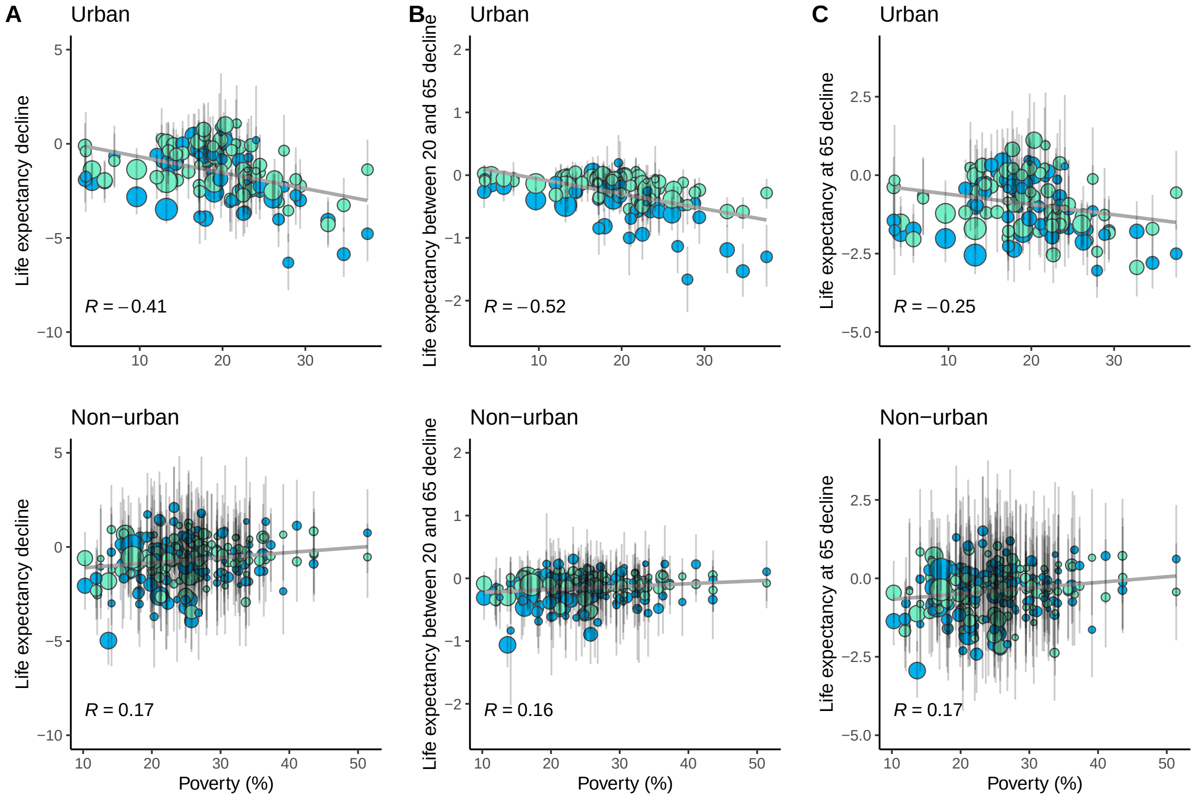


Figure 14: Declines in life expectancy at birth (A), life expectancy between 20 and 65 (B), and life expectancy at 65 (C) as a function of proportion of population that lives in poverty. Each dot is a municipality, separated by gender (colors) Urban and non-urban municipalities are shown in first and second row, respectively. A strong effect appears in urban setups, and the correlation is stronger in for life expectancy between 20 and 65.

Figure 15: Declines in life expectancy at birth (A), life expectancy between 20 and 65 (B), and life expectancy at 65 (C) as a function of proportion of population that lives in a crowded home. Each dot is a municipality, separated by gender(colors) Urban and non-urban municipalities are shown in first and second row, respectively.

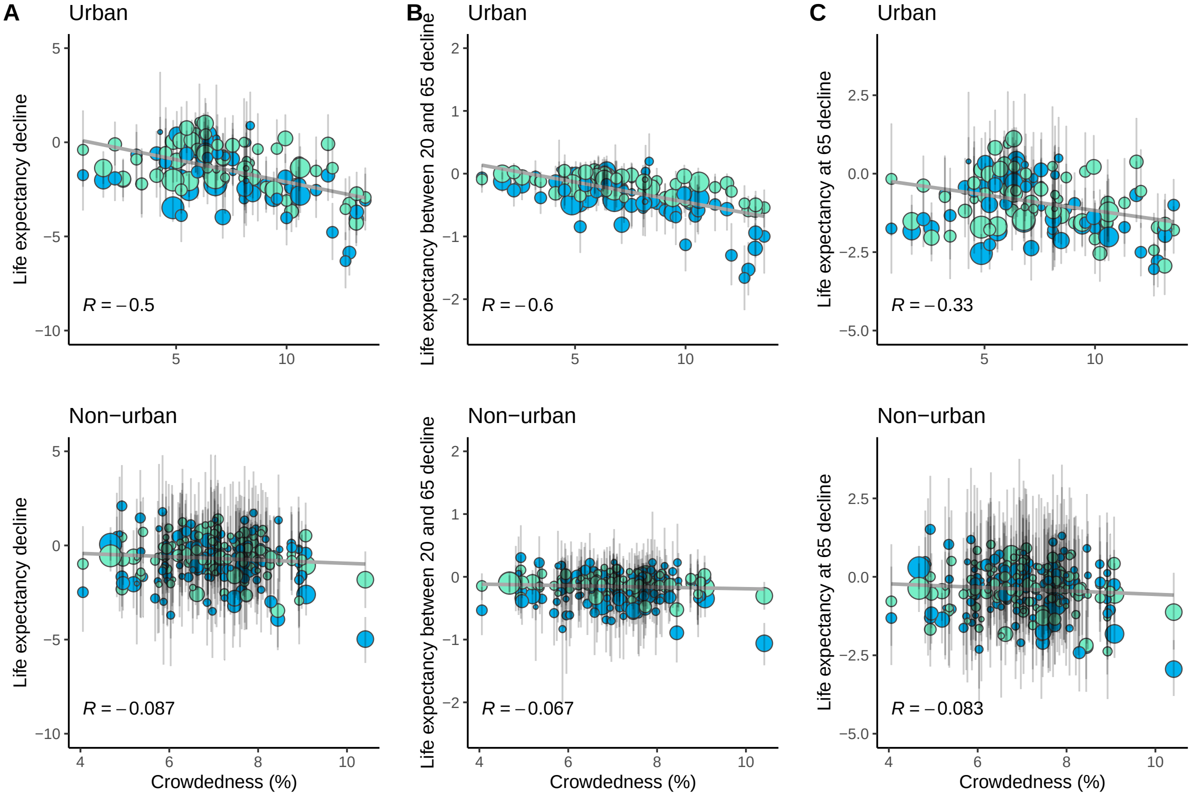

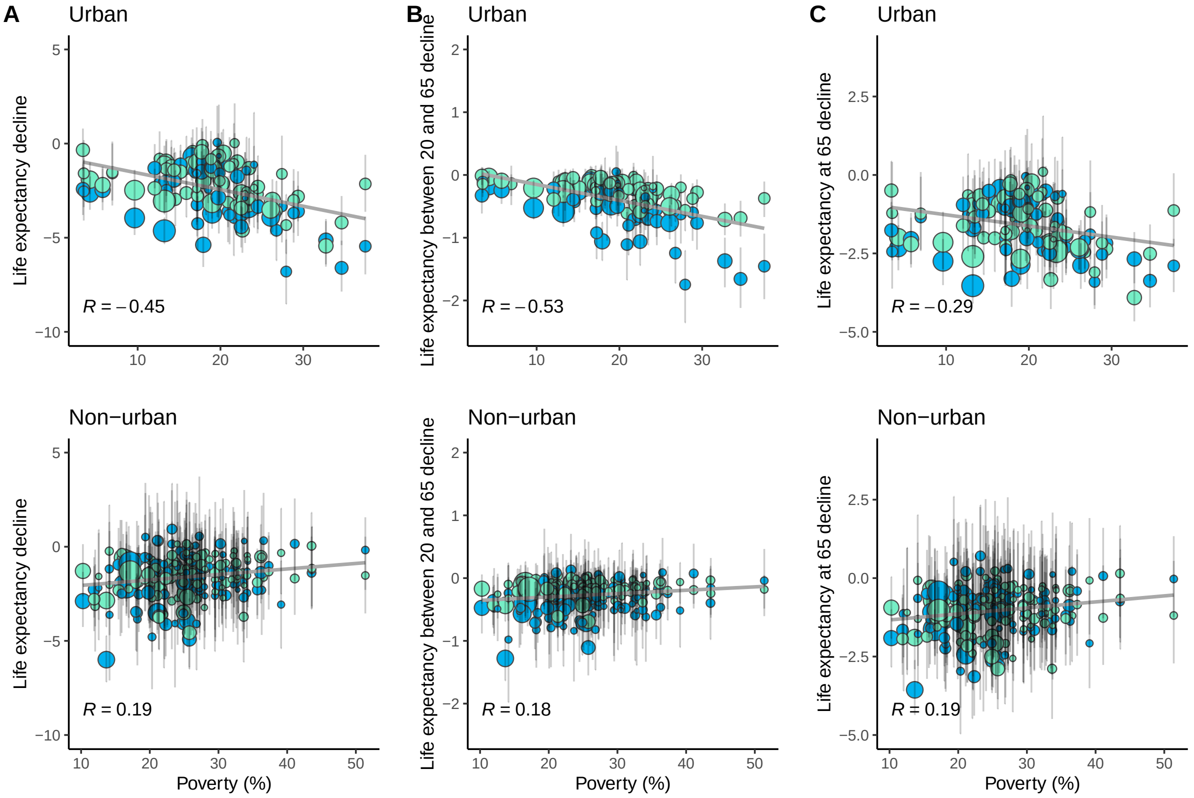


Figure 16: Same as 14 but with population estimates for years 2017,2018,2019,2020 all equal to population counts in 2017 as given by census.

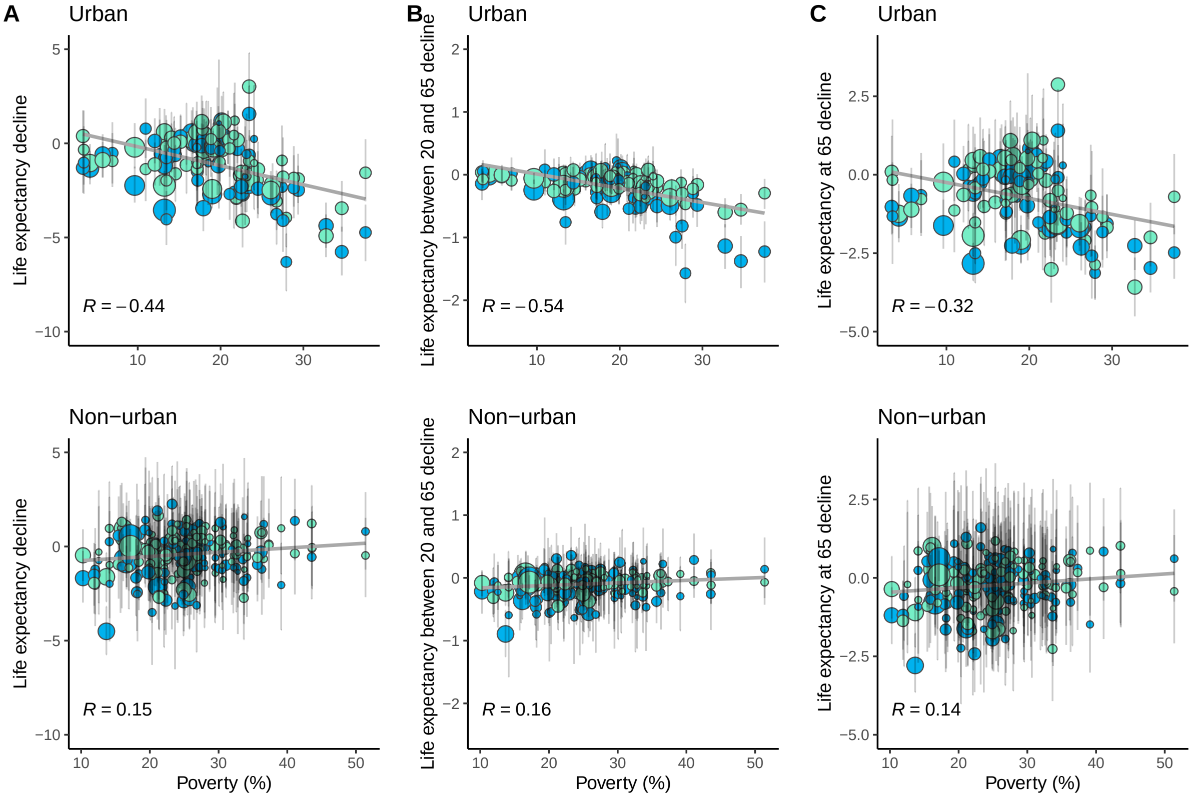


Figure 17: Same as 14 but with population estimates given by official projections.
